# Comparison and trends in outcomes of inborn and outborn infants born before 33 weeks’ gestation

**DOI:** 10.1101/2025.06.04.25329014

**Authors:** Gergely Balazs, Melinda Vojtko, Mariann Marki, Samuel Lowe, Magdolna Riszter, Gusztav Belteki, Andras Balajthy

**Affiliations:** Division of Neonatology, Department of Pediatrics, Faculty of Medicine, University of Debrecen, Debrecen, Hungary; Addenbrooke’s Hospital, Cambridge University Hospitals NHS Foundation Trust, Cambridge, United Kingdom; Neonatal Intensive Care Unit, Cambridge University Hospitals NHS Foundation Trust, Cambridge, United Kingdom

## Abstract

**Objective:** To compare clinical care and outcome of preterm infants born in a tertiary neonatal unit with those requiring early postnatal transport to the same unit, and analyze their trends over an 9-year period.

**Study Design:** A retrospective study of infants born before 33 weeks’ gestation between 2013 and 2021. Inborn (n=913) and outborn (n=133) infants were compared using logistic regression and trend analysis.

**Result:** Outborn infants more frequently required intubation in delivery room (59.4% vs. 32.3%, p<0.001) and mechanical ventilation (69.2% vs. 44.5 %, p<0.001). They more frequently had severe intraventricular haemorrhage (IVH, 15% vs. 8.1%, p=0.027) and had lower survival rate (88.7% vs. 91.9%, p=0.02). Birth outside a tertiary neonatal unit increased the risk of severe IVH [odds ratio: 2.4 (95% confidence interval: 1.3-4.3)]. Only inborn infants showed decreasing trends in delivery room intubation (annual percentage change, APC: -21.2%, p<0.001) and mechanical ventilation (APC: -8.3%, p=0.001).

**Conclusion:** Outborn infants continue to require more invasive respiratory support and experience worse outcomes.

## Introduction

Although premature infants born before the 33^rd^ gestational week constitute only 1-2% of all neonates, the resources their care requires pose significant challenges to healthcare facilities (1). Due to advances in prenatal and postnatal interventions, mortality of this population has decreased globally (2). However, the risk of major morbidities such as intraventricular haemorrhage (IVH), bronchopulmonary dysplasia (BPD), necrotizing enterocolitis (NEC), retinopathy of prematurity (ROP), periventricular leukomalacia (PVL) has remained substantial in this group (1).

Centralization of neonatal intensive care can significantly improve the short- and long-term outcomes of preterm infants; however, *in utero* transport is not always possible or feasible (3). Over the last two decades, care during postnatal transport has changed significantly, and mortality as well as morbidity of outborn preterm infants have improved (4,5). However, large comparative studies suggest that outcome of preterm infants born in hospitals with tertiary neonatal intensive care units (NICUs) has remained significantly better than of those born outside these centres (3, 6-10). Among the distinct adverse outcomes, most studies highlighted the association between early postnatal transport and severe brain injury (6,10-12).

Comparing clinical care and outcome of preterm infants born within and outside a tertiary centre can facilitate further development of local guidelines and procedures and monitoring their impact, thereby improving clinical practice, resource allocation and, potentially, clinical outcomes. However, the long data collection period usually required due to the relatively low number of outborn patients and the necessarily retrospective study design together may make it difficult to assess the impact of newly implemented guidelines and interventions for this group of patients (7,8,10). An alternative approach is employing trend analytical methods, which may circumvent some of these difficulties (5,13).

In this study we compared respiratory care, outcome and their temporal trends over an 9-year period of infants born before the completed 33 weeks of gestation in a hospital with a tertiary neonatal intensive care unit and infants who required early postnatal transport to the same unit. We hypothesized that morbidity and mortality of outborn preterm infants will be significantly worse than those of the inborn population.

## Methods

### Patients

We retrospectively collected and analyzed data from preterm infants born before 33 completed gestational weeks, who were admitted to the regional tertiary NICU of the University of Debrecen Clinical Centre over a 9-year period (2013–2021). Infants were either delivered in the same building (“inborn infants”) or transferred postnatally within the first 72 hours of life (“outborn infants”). The regional neonatal transport service of the University of Debrecen Clinical Centre covers an area with an approximate radius of 130 kilometers. In addition to the Clinical Centre itself, the service includes five district general hospitals with level 1 or 2 neonatal units. A dedicated neonatal transport team is available 24/7 to ensure the emergency transport of preterm infants. The transport team consists of a fully trained neonatologist experienced in neonatal transport and an experienced neonatal transport nurse practitioner. In line with international standards, *in utero* transport is prioritized and very preterm deliveries in the local hospitals occur only in emergency situations when inter-hospital transport of the mother is not possible. In cases of threatened preterm birth, referring institutions immediately notify the regional unit and the neonatal transport service, and the transport team attends these deliveries as time-critical emergencies with blue lights and ambulance priorities. However, due to the distances involved, initial delivery room care is provided by the staff of the referring hospital, according to the relevant national guidelines, with the transport team joining them upon arrival. Care during transport was provided by the neonatal transport team following standard operating procedures of the team.

### Data collection and outcome variables

The study was approved by the Regional and Institutional Research Ethics Committee of the University of Debrecen. Data were collected from handwritten and electronic neonatal and maternal health care records and transport forms. The authors de-identified all patients’ information during extraction. Data collection was completed between November and December 2022. The regional NICU is affiliated with the Vermont-Oxford Network, and utilizes the “Nightingale Data Definitions” guide for clinical definitions, consistent with their routine practice (14). BPD was defined as the need of supplemental oxygen at 36 weeks of postmenstrual age (PMA) or having been discharged on oxygen treatment before 36 weeks of PMA. IVH was assessed by cranial ultrasonography and graded according to Papile *et al* (15). The term “severe IVH” was used for grade ≥3 IVH. ROP was defined using the International Classification for Retinopathy of Prematurity and referred to as “severe” if it was stage ≥3 (16). Periventricular leukomalacia (PVL) was defined as presence of one or more cysts in the periventricular region of the brain, detected by serial ultrasound scans. Late onset sepsis (LOS) was defined as a culture of a pathogenic organism (bacterium or fungus) from blood and/or cerebrospinal fluid after 72 hours of age. Surgical necrotizing enterocolitis (NEC) was defined as NEC requiring a surgical intervention (i.e., laparotomy, laparoscopy, intraperitoneal drain) due to intestinal perforation or failure to improve with medical management. Mortality was defined as death before discharge from hospital or death before one year of age in case of prolonged hospitalization. Morbidity-free survival was defined as the lack of BPD, NEC, PVL, LOS and severe IVH up to the time of discharge from hospital. ROP was not included due to the continuation of follow-up care beyond hospital discharge.

### Data analysis

Descriptive statistics for the study population included mean and standard deviation or median and interquartile range for continuous variables, and count and percentage for categorical variables. Comparison of variables between the two groups was performed using Student’s t-test, Mann-Whitney U-test, and Chi-square tests, depending on the type and distribution of the data. The impact of outborn status on the outcomes of preterm infants, considering other known demographic or perinatal risk factors (gestational age, gender, presence or absence of intrauterine growth restriction, IUGR, presence or absence of antenatal steroids and mode of delivery), was determined using binary logistic regression. Antenatal steroid prophylaxis (ANS) was considered administered if at least one dose was given prior to delivery. Differences were considered statistically significant at p<0.05. Statistical analysis was performed using the SPSS V25 program. Trends over the years and changes in trends were examined using the Joinpoint Regression Program (the Surveillance Research Program of the National Cancer Institute, Bethesda, MD, USA). The Joinpoint Regression Program determines breakpoints and selects the best-fitting model among log-linear regression models fitted to the data points using Monte Carlo permutation (5,17). Changes in trends are characterized by the annual percentage change (APC). The program also allows for comparing trends between patient groups. If trends run parallel, there is no significant difference in APC values.

## Results

### Outborn infants received antenatal steroids less frequently and required mechanical ventilation more frequently than inborn infants

A total of 913 inborn and 133 outborn preterm infants were included in the study. Although the number of cases in the two groups differed considerably, the annual ratio of the two study groups did not change significantly over the study period (APC = -0.58, p=0.874). Gestational age, birth weight and gender distribution were balanced throughout the study period; however, outborn infants had significantly lower 1- and 5-minute Apgar scores (Table 1). Inborn preterm infants were more likely to receive ANS and to be delivered by Caesarean section. Outborn infants were more frequently intubated in the delivery room and more of them required mechanical ventilation during their stay on NICU (Table 1).

**Table 1:** Comparison of demographic and clinical characteristics between the two study populations.

|  | Inborn (n=913) | Outborn (n=133) | p-value |
| --- | --- | --- | --- |
| Gestational age (weeks), median (IQR<sup>a</sup>) | 28 (26-30) | 28 (26-30) | 0.18 |
| Birth weight (g), median (IQR) | 980 (730- 1340) | 1100 (805-1310) | 0.10 |
| Male, n (%) | 504 (55.2) | 71 (53).4 | 0.69 |
| 1-minute Apgar score, median (IQR) | 7 (5-8) | 5 (3-7) | <0.001 |
| 5-minute Apgar score, median (IQR) | 8 (7-8) | 7 (5-8) | <0.001 |
| Admission temperature (C°), mean (SD<sup>b</sup>) | 35.76 (0.73) | 35.37 (1.42) | 0.54 |
| Caesarean section, n (%) | 679 (74.4) | 77 (57.9) | <0.001 |
| Antenatal steroid, n (%) | 833 (91.2) | 55 (41.4) | <0.001 |
| Delivery room intubation, n (%) | 295 (32.3) | 79 (59.4) | <0.001 |
| Surfactant therapy, n (%) | 591 (64.7) | 102 (76.7) | 0.006 |
| Mechanical ventilation, n (%) | 406 (44.5) | 92 (69.2) | <0.001 |

<sup>a</sup>IQ: Interquartile range, the range between the 75<sup>th</sup> and 25<sup>th</sup> percentiles.
<sup>b</sup>SD: standard deviation.

### Outcome of inborn and outborn infants

Outborn infants had significantly lower survival to discharge and lower morbidity-free survival rate (Table 2). Severe IVH and LOS occurred more frequently in outborn infants. There was no significant difference in the other morbidities.

**Table 2:**
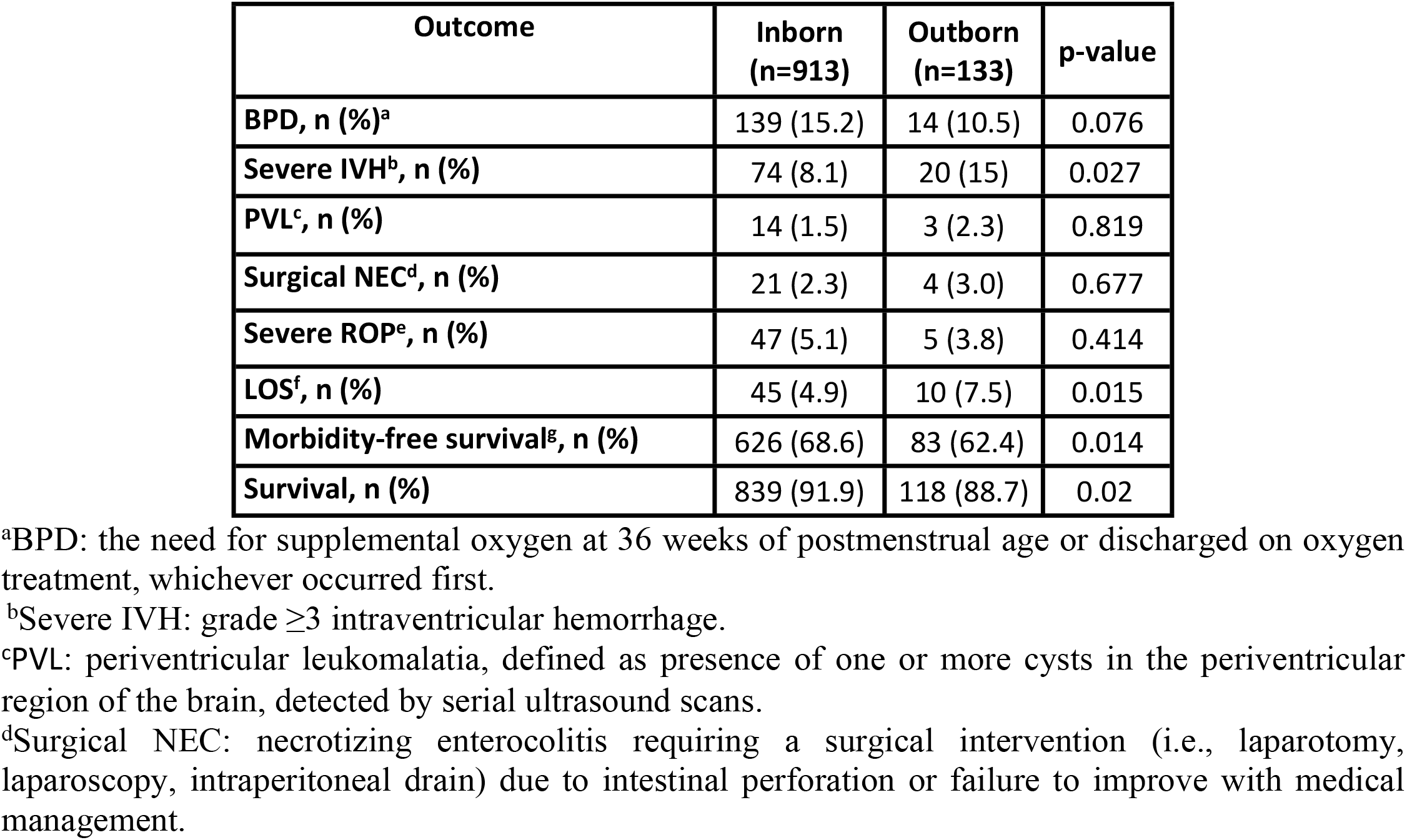
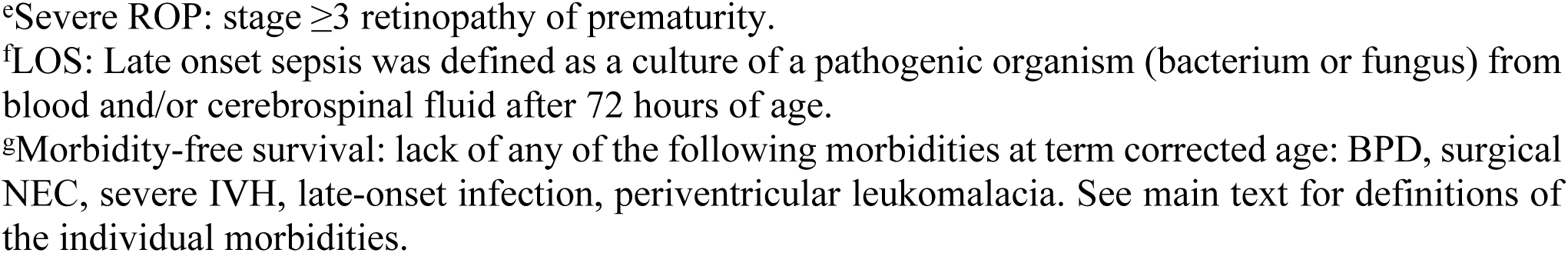
Comparing neonatal outcomes of inborn and outborn infants at term corrected age.

| Outcome | Inborn (n=913) | Outborn (n=133) | p-value |
| --- | --- | --- | --- |
| BPD, n (%)<sup>a</sup> | 139 (15.2) | 14 (10.5) | 0.076 |
| Severe IVH<sup>b</sup>, n (%) | 74 (8.1) | 20 (15) | 0.027 |
| PVL<sup>c</sup>, n (%) | 14 (1.5) | 3 (2.3) | 0.819 |
| Surgical NEC<sup>d</sup>, n (%) | 21 (2.3) | 4 (3.0) | 0.677 |
| Severe ROP<sup>e</sup>, n (%) | 47 (5.1) | 5 (3.8) | 0.414 |
| LOS<sup>f</sup>, n (%) | 45 (4.9) | 10 (7.5) | 0.015 |
| Morbidity-free survival<sup>g</sup>, n (%) | 626 (68.6) | 83 (62.4) | 0.014 |
| Survival, n (%) | 839 (91.9) | 118 (88.7) | 0.02 |

<sup>a</sup>BPD: the need for supplemental oxygen at 36 weeks of postmenstrual age or discharged on oxygen treatment, whichever occurred first.
<sup>b</sup>Severe IVH: grade $\geq 3$ intraventricular hemorrhage.
<sup>c</sup>PVL: periventricular leukomalacia, defined as presence of one or more cysts in the periventricular region of the brain, detected by serial ultrasound scans.
<sup>d</sup>Surgical NEC: necrotizing enterocolitis requiring a surgical intervention (i.e., laparotomy, laparoscopy, intraperitoneal drain) due to intestinal perforation or failure to improve with medical management.
<sup>e</sup>Severe ROP: stage $\geq 3$ retinopathy of prematurity.
<sup>f</sup>LOS: Late onset sepsis was defined as a culture of a pathogenic organism (bacterium or fungus) from blood and/or cerebrospinal fluid after 72 hours of age.
<sup>g</sup>Morbidity-free survival: lack of any of the following morbidities at term corrected age: BPD, surgical NEC, severe IVH, late-onset infection, periventricular leukomalacia. See main text for definitions of the individual morbidities.

After adjusting for known demographic and perinatal risk factors in a logistic regression model, outborn status remained significantly associated with mechanical ventilation but there was no significant difference between the inborn and the outborn group in survival or in morbidities (Table 3).

**Table 3:**
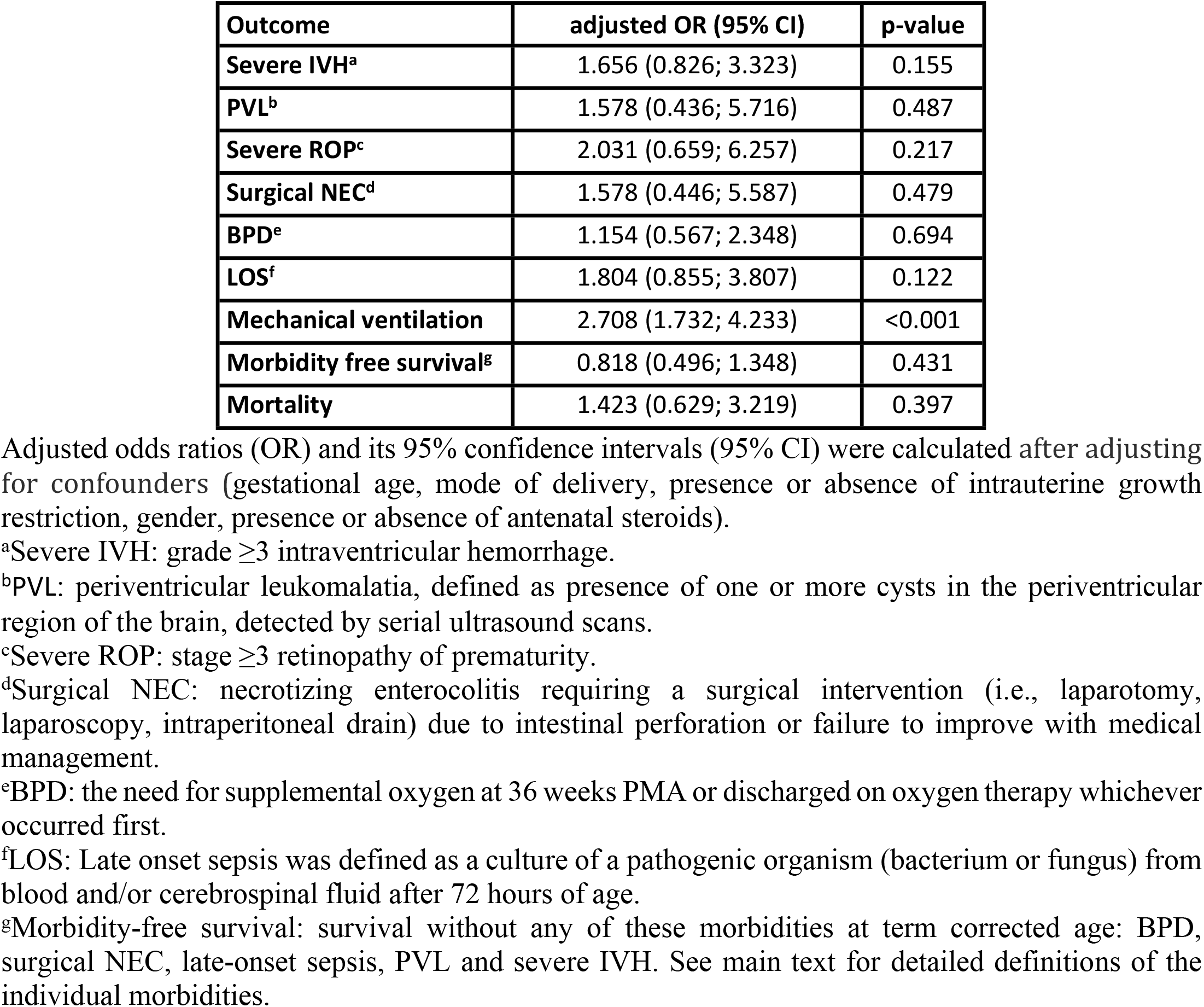
Analysing the influence of outborn status on neonatal outcomes using multivariable logistic regression models1.

### Decreasing trends in delivery room intubation and mechanical ventilation in inborn but not in outborn infants

We also analysed temporal trends over the study period. The frequency of Caesarean section, antenatal steroid prophylaxis, and surfactant therapy did not change significantly over the study period (Table 4). For inborn preterm infants, there was a significantly decreasing trend in delivery room intubation and in the need for mechanical ventilation from 2013 to 2021, while there was no change in these parameters among postnatally transported infants (see Fig 1 A and B). Decreasing trends were observed in all morbidities, but they were not statistically significant (Fig 1 C-F). Overall and morbidity-free survival showed a non-significant increasing trend in both patient groups (Fig 1 G-H, Table 4).

**Table 4:** Comparing trends of neonatal care and outcome by Joinpoint regression analysis.

|  | Inborn |  | Outborn |  | p<sub>APC</sub> |
| --- | --- | --- | --- | --- | --- |
|  | APC | p | APC | p |  |
| Caesarean section | -0.35 | 0.637 | -0.35 | 0.637 | 0.07 |
| Antenatal steroid | -0.62 | 0.16 | -0.62 | 0.16 | 0.94 |
| Mechanical ventilation | -8.3 | 0.001 | -0.4 | 0.9 | 0.04 |
| Surfactant therapy | -2.77 | 0.028 | -2.77 | 0.028 | 0.39 |
| Delivery room intubation | -21.2 | <0.001 | 0 | 0.99 | 0.004 |
| BPD<sup>a</sup> | -6.5 | 0.175 | -6.5 | 0.175 | 0.28 |
| Severe IVH<sup>b</sup> | -7.9 | 0.13 | -7.9 | 0.13 | 0.7 |
| PVL<sup>c</sup> | -3.4 | 0.457 | -3.4 | 0.457 | 0.52 |
| Surgical NEC<sup>d</sup> | -3.4 | 0.54 | -3.4 | 0.54 | 0.56 |
| Severe ROP<sup>e</sup> | -4.9 | 0.43 | -4.9 | 0.43 | 0.22 |
| LOS<sup>f</sup> | 8.8 | 0.08 | 8.8 | 0.08 | 0.70 |
| Morbidity-free survival<sup>g</sup> | 1.8 | 0.24 | 1.8 | 0.24 | 0.3 |
| Survival | 0.2 | 0.69 | 0.2 | 0.69 | 0.82 |

Changes in trends are characterized by the annual percentage change (APC) for each patient group.
p<sub>APC</sub> represents p-value of the statistical test of parallelism (if trends run parallel p<sub>APC</sub>>0.05).
<sup>a</sup>BPD: the need for supplemental oxygen at 36 weeks PMA or discharged on oxygen treatment whichever occurred first.
<sup>b</sup>Severe IVH: grade $\geq 3$ intraventricular hemorrhage.
<sup>c</sup>PVL: periventricular leukomalacia, defined as presence of one or more cysts in the periventricular region of the brain, detected by serial ultrasound scans.
<sup>d</sup>Surgical NEC: necrotizing enterocolitis requiring a surgical intervention (i.e., laparotomy, laparoscopy, intraperitoneal drain) due to intestinal perforation or failure to improve with medical management.
<sup>e</sup>Severe ROP: stage $\geq 3$ retinopathy of prematurity.
<sup>f</sup>LOS: Late onset sepsis was defined as a culture of a pathogenic organism (bacterium or fungus) from blood and/or cerebrospinal fluid after 72 hours of age.
<sup>g</sup>Morbidity-free survival: lack of any of the following morbidities at term corrected age: severe IVH, BPD, surgical NEC, pneumothorax requiring chest drain, culture positive late-onset infection, periventricular leukomalacia. See main text for definitions of the individual morbidities

**Fig 1.**
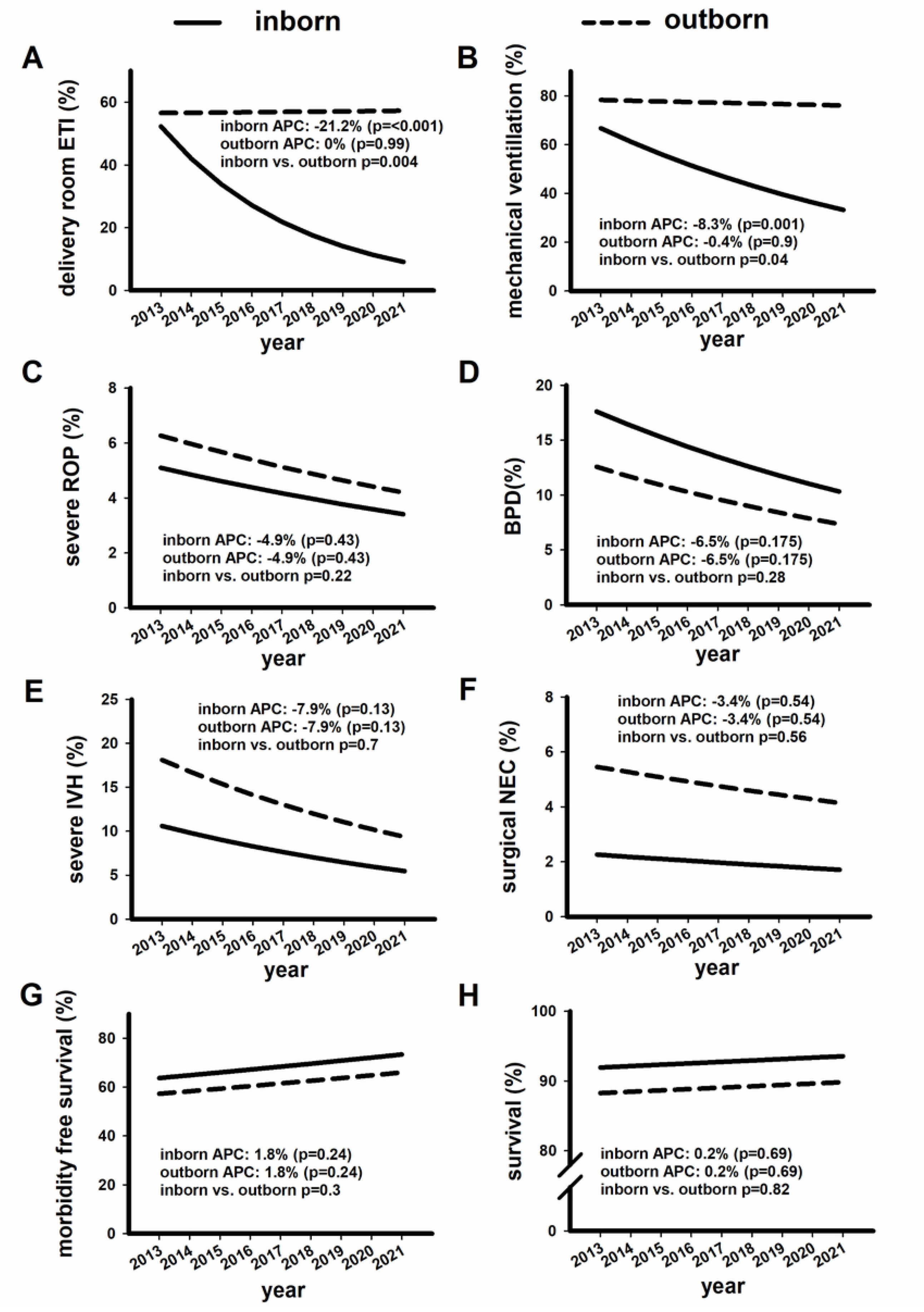
Trends in respiratory care and outcomes of inborn and outborn infants. Trends in delivery room intubation (A), mechanical ventilation (B), severe ROP (C), BPD (D), severe IVH (E), surgical NEC (F), morbidity-free survival (G), and survival (H) in inborn and outborn infants over the study period. Annual percentage change (APC) values were used to characterize the trend curves separately for both groups. A p-value <0.05 indicates a statistically significant change in APC. We also compared the APC values of inborn and outborn neonates for each variable; a p-value <0.05 was considered a statistically significant difference.

## Discussion

In this study, we compared early care and outcome of inborn and outborn preterm infants born before 33 weeks of gestation in a large tertiary perinatal centre over an 9-year period. During the study period, the proportion of inborn infants in the region covered by our tertiary NICU was relatively high (87%), similar to other developed regions (7, 18-21). There was no significant improvement in *in utero* transfer rate of our region over the study period. In a cohort study by Lecour *et al.* with comparable outborn rate, birth outside tertiary centres could have been prevented only in 9.6% of cases (21). In order to further reduce the proportion of cases requiring postnatal transport, it is essential to regularly review current recommendations and practices, monitor case numbers, and to ensure continuous communication between referring and receiving units at a regional level (7,22,23).

Antenatal steroid prophylaxis significantly improves outcomes in preterm infants (24). We demonstrated that the rate of antenatal steroid prophylaxis was significantly lower in the outborn group, consistent with previous studies (8,17). There was no significant change in the trend of steroid prophylaxis in either group over the study period. Considering the high *in utero* transport rate, it is likely that the outborn group represents cases with rapidly progressing labor and maternal or fetal conditions necessitating immediate delivery. This suggests that it will be difficult to make a significant improvement in steroid exposure of the outborn group in the future. Of note, antenatal steroid prophylaxis, even if given only hours before delivery, may improve outcome (25).

In line with international recommendations and trends, the percentage of infants intubated in delivery room decreased each year for the inborn group, and an increasing number of them was stabilized with non-invasive respiratory support (26,27). There was no improvement in these indicators in the outborn group over the study period. Moreover, outborn infants received mechanical ventilation more frequently even after accounting for differences in demographic and perinatal factors. This may be due to limited experience in district hospitals, worse general condition of outborn infants (e.g., lack of steroid prophylaxis, lower 1-minute Apgar score), or lower threshold for intubating them to ensure a safe postnatal transfer.

The incidence of severe IVH was found to be significantly higher in patients requiring postnatal transport in several studies (6,9,10,28). The cause of this association is not fully understood, but its mechanism is likely to be multifactorial, including less experienced staff attending deliveries, transport’s physical forces, temperature instability, higher incidence of mechanical ventilation and lower steroid prophylaxis rate (3,6,29-31). In our cohort, outborn infants also more frequently had severe IVH; however, the difference was not significant once demographic and antenatal factors have been accounted for in a logistic regression model. Since demographic factors (e.g., gestation, birth weight, gender) were not significantly different between the group, we think that the higher severe IVH rate in our outborn cohort was to a large extent due to their lower antenatal steriod exposure, and this in turn may have contributed to the lower overall and mobidity-free survival among them. In another retrospective cohort study, a full course of antenatal steroid prophylaxis significantly decreased the risk of IVH irrespective of transport status, but even after a full course of steroid prophylaxis, transferred outborn infants born at less than 28 weeks of gestation were still more likely to develop IVH than their inborn counterparts (10). Of note, the IVH rate can be affected by factors difficult to include in regression models such as details of neonatal service delivery details of the region.

We also observed a higher rate of LOS in the outborn group, in alignment with previous studies (32). The increased frequency of delivery room intubation and mechanical ventilation in this group may contribute to a higher incidence of bacterial infections (33).

Our study has several limitations. First, we were not able to collect information on premature infants who died in the referring institutions’ delivery rooms, so only outborn infants transferred to the tertiary centre were included in the analysis (of the inborn infants, three died in the delivery room). Therefore, the difference in mortality between inborn and outborn infants may have been higher than we have reported. Second, for rarely occurring complications (ROP, NEC), the low case numbers could have significantly distorted the relevant results. Third, generalizability of the results may be affected by specificities of neonatal care delivery within our neonatal network.

## Conclusion

Delivery room intubation and mechanical ventilation have shown decreasing trends over the last decade for inborn but not for outborn infants. Mechanical ventilation, severe IVH and late-onset sepsis are more frequent in outborn infants.

## Data Availability

All relevant data are within the manuscript and its Supporting Information files.

## Ethical approval

The study was approved by the Scientific and Research Ethics Committee of the University of Debrecen under the registration number of „DE RKEB/IKEB 6189-2022” and was conducted in accordance with the ethical standards of all relevant national and institutional committees and the World Medical Association’s Declaration of Helsinki. For this retrospective study, informed consent was waived as sanctioned.

